# Time in treatment: Examining mental illness trajectories across inpatient psychiatric treatment

**DOI:** 10.1101/2020.06.26.20140293

**Authors:** Hyuntaek Oh, Jaehoon Lee, Seungman Kim, Katrina A. Rufino, Peter Fonagy, John M. Oldham, Bella Schanzer, Michelle A. Patriquin

## Abstract

Early discharge or reduced length of stay for inpatient psychiatric patients is related to increased readmission rates and worse clinical outcomes including increased risk for suicide. Trajectories of mental illness outcomes have been identified as an important method for predicting the optimal length of stay but the distinguishing factors that separate trajectories remain unclear. We sought to identify the distinct classes of patients who demonstrated similar trajectories of mental illness over the course of inpatient treatment, and we explore the patient characteristics associated with these mental illness trajectories. We used data (*N* = 3,406) from an inpatient psychiatric hospital with intermediate lengths of stay. Using growth mixture modeling, latent mental illness scores were derived from six mental illness indicators: psychological flexibility, emotion regulation problems, anxiety, depression, suicidal ideation, and disability. The patients were grouped into three distinct trajectory classes: (1) High-Risk, Rapid Improvement (HR-RI); (2) Low-Risk, Gradual Improvement (LR-GI); and (3) High-Risk, Gradual Improvement (HR-GI). The HR-GI was significantly younger than the other two classes. The HR-GI had significantly more female patients than males, while the LR-GI had more male patients than females. Our findings indicated that younger females had more severe mental illness at admission and only gradual improvement during the inpatient treatment period, and they remained in treatment for longer lengths of stay, than older males.

## Introduction

Inpatient psychiatry lengths of stay (LOS) in the United States have been becoming shorter and shorter, with the average inpatient LOS now approximately five to six days (Glick et al., 2011; Sturm and Bao, 2000). Although some studies indicate that shorter LOS is as effective as longer LOS (e.g., 30 days) in reducing symptoms of dementia (Kunik et al., 2001), major depression (Pettit et al., 2005), and schizophrenia (Johnstone and Zolese, 1999), others have indicated that psychiatric morbidity is greater at discharge among psychiatric patients with a shorter LOS than those with a longer LOS (Glick et al., 2011; Ruaño et al., 2013). Meta-analytic results confirm that patients with a history of psychiatric disorders are at an increased risk of suicide (Harris and Barraclough, 1997) and the suicide risk is particularly high when the patients were discharged from psychiatric hospitals where the average LOS is between 5 to 7 days (Appleby et al., 1999; Goldacre et al., 1993). This is congruent with the literature indicating that a LOS of less than 14 days in inpatient psychiatry was significantly associated with a higher suicide risk (Desai et al., 2005). In addition to increased suicide risk with reduced LOS, other studies demonstrated that shorter LOS predicts greater risk of readmission (Lin et al., 2006; Ruaño et al., 2013). Despite the relationship between poorer outcome and reduced LOS, economic pressures to limit LOS for inpatient hospitalization have tended to tip the scale in favor of less time being needed for inpatient treatment (Compton et al., 2006; Hirsch et al., 1979).

In addition to economic pressure, studies have identified other clinical factors which appear to influence LOS for inpatient psychiatry. These include: prior hospitalization (Stevens et al., 2001), level of psychopathology (Cohen and Casimir, 1989), medical comorbidity (Lyketsos et al., 2002), suicide risk (Cohen and Casimir, 1989), substance use (Warnke and Rossler, 2008), and psychiatric diagnoses (Blader, 2011). Hospitalization history and diagnoses of schizophrenia, major depression, and other psychotic disorders were correlated with extended LOS, whereas substance use was associated with reduced LOS (Blader, 2011; Clapp et al., 2013; Cohen and Casimir, 1989; Hallak et al., 2003; Stevens et al., 2001; Warnke and Rossler, 2008). Associations between LOS and demographic information (e.g., sex, marital status, homelessness) were also noted in several studies but findings are mixed (Averill et al., 2001; Cohen and Casimir, 1989; Warnke and Rossler, 2008). A limitation of the correlational approach taken to identifying predictors of LOS is that it assumes an underlying homogeneity and explores primarily linear predictors of LOS. In principle it is perfectly possible that in some instances, when it retards recovery severity predicts increased LOS while in others, where high severity scores expedite rapid improvement, the association is reversed. Notably, there have been few studies that examined the trajectories of recovery over the course of psychiatric hospitalization and how these patterns can be predicted from patient characteristics, and how they relate to treatment and follow-up outcomes. Being able to identify groups of patients based on information available on admission (e.g. demographic, self-reported symptom data), allows clinicians to anticipate likely treatment response and support effective decision making by stratifying care (Saunders et al., 2019).

Growth mixture modeling (GMM) is a statistical method designed to identify classes of individuals who are homogeneous in terms of their longitudinal change in a set of variables of interest (Muthén, 2006; Ram and Grimm, 2009). GMM has been useful for recognizing classes of patients who share similar longitudinal trajectories of depressive symptoms (Bombardier et al., 2016; Gomez et al., 2017; Saunders et al., 2019; Saunders et al., 2020). However, these were either studies of outpatient treatments or did not consider other psychiatric disorders or their comorbidity. Another study, although the finding could be limited to older adults, has also utilized GMM to examine the factors related to longitudinal trajectories of PTSD symptoms after a hurricane disaster (Pietrzak et al., 2013).

The present study applied GMM to longitudinal clinical data obtained from a large sample of adults admitted to an inpatient psychiatric hospital. We considered six key psychiatric indicators to agnostically determine trajectories of change in mental illness outcomes during inpatient psychiatric treatment: psychological flexibility (Kashdan and Rottenberg, 2010), emotion regulation problems (Gross and Muñoz, 1995), anxiety (Ormel et al., 1994), depression (Ormel et al., 1994), suicide ideation severity (Kessler et al., 1999), and disability (Ormel et al., 1994). The aims of this study were to (1) establish the patterns of change in mental health, and (2) identify the variables associated with the trajectories. It was hypothesized that different mental illness trajectories would emerge (e.g., those that showed improvement, those that showed treatment resistance), yet the specific number of trajectories was not assumed as this was considered an exploratory approach. We also examined a number of demographic and clinical variables (see Table 1 and 2) that are relevant to mental illness outcomes and hospitalization to identify the demographic and clinical variables associated with the identified patterns of change in mental illness. Lastly, we hypothesized that improved mental illness outcomes would be associated with longer LOS, rather than shorter LOS.

**Table 1.**
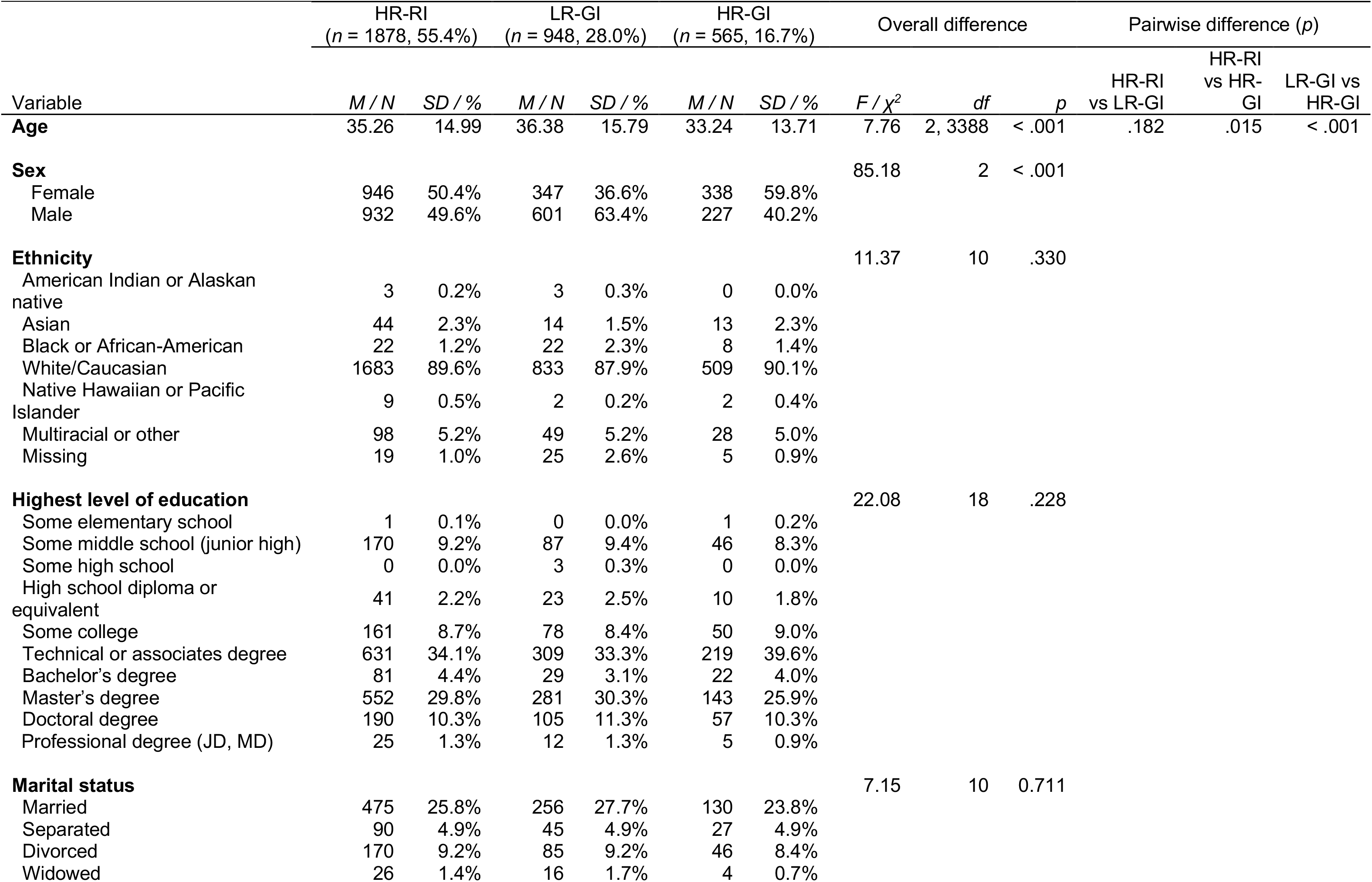
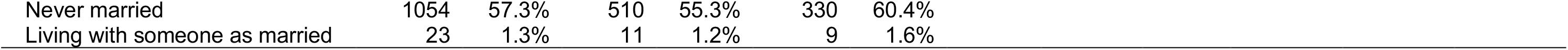
Comparisons of demographic variables between the classes. HR-RI: High-risk, Rapid improvement, LR-GI: Low-risk, Gradual improvement, HR-GI: High-risk, Gradual improvement, M: mean, SD: standard deviation.

| Variable | HR-RI<br>(n = 1878, 55.4%) |  | LR-GI<br>(n = 948, 28.0%) |  | HR-GI<br>(n = 565, 16.7%) |  | Overall difference |  |  | Pairwise difference (p) |  |  |
| --- | --- | --- | --- | --- | --- | --- | --- | --- | --- | --- | --- | --- |
| | M / N | SD / % | M / N | SD / % | M / N | SD / % | F / $\chi^2$ | df | p | HR-RI<br>vs LR-GI | HR-RI<br>vs HR-GI | LR-GI vs<br>HR-GI |
| **Age** | 35.26 | 14.99 | 36.38 | 15.79 | 33.24 | 13.71 | 7.76 | 2, 3388 | < .001 | .182 | .015 | < .001 |
| **Sex** |  |  |  |  |  |  | 85.18 | 2 | < .001 |  |  |  |
| Female | 946 | 50.4% | 347 | 36.6% | 338 | 59.8% |  |  |  |  |  |  |
| Male | 932 | 49.6% | 601 | 63.4% | 227 | 40.2% |  |  |  |  |  |  |
| **Ethnicity** |  |  |  |  |  |  | 11.37 | 10 | .330 |  |  |  |
| American Indian or Alaskan<br>native | 3 | 0.2% | 3 | 0.3% | 0 | 0.0% |  |  |  |  |  |  |
| Asian | 44 | 2.3% | 14 | 1.5% | 13 | 2.3% |  |  |  |  |  |  |
| Black or African-American | 22 | 1.2% | 22 | 2.3% | 8 | 1.4% |  |  |  |  |  |  |
| White/Caucasian | 1683 | 89.6% | 833 | 87.9% | 509 | 90.1% |  |  |  |  |  |  |
| Native Hawaiian or Pacific<br>Islander | 9 | 0.5% | 2 | 0.2% | 2 | 0.4% |  |  |  |  |  |  |
| Multiracial or other | 98 | 5.2% | 49 | 5.2% | 28 | 5.0% |  |  |  |  |  |  |
| Missing | 19 | 1.0% | 25 | 2.6% | 5 | 0.9% |  |  |  |  |  |  |
| **Highest level of education** |  |  |  |  |  |  | 22.08 | 18 | .228 |  |  |  |
| Some elementary school | 1 | 0.1% | 0 | 0.0% | 1 | 0.2% |  |  |  |  |  |  |
| Some middle school (junior high) | 170 | 9.2% | 87 | 9.4% | 46 | 8.3% |  |  |  |  |  |  |
| Some high school | 0 | 0.0% | 3 | 0.3% | 0 | 0.0% |  |  |  |  |  |  |
| High school diploma or<br>equivalent | 41 | 2.2% | 23 | 2.5% | 10 | 1.8% |  |  |  |  |  |  |
| Some college | 161 | 8.7% | 78 | 8.4% | 50 | 9.0% |  |  |  |  |  |  |
| Technical or associates degree | 631 | 34.1% | 309 | 33.3% | 219 | 39.6% |  |  |  |  |  |  |
| Bachelor's degree | 81 | 4.4% | 29 | 3.1% | 22 | 4.0% |  |  |  |  |  |  |
| Master's degree | 552 | 29.8% | 281 | 30.3% | 143 | 25.9% |  |  |  |  |  |  |
| Doctoral degree | 190 | 10.3% | 105 | 11.3% | 57 | 10.3% |  |  |  |  |  |  |
| Professional degree (JD, MD) | 25 | 1.3% | 12 | 1.3% | 5 | 0.9% |  |  |  |  |  |  |
| **Marital status** |  |  |  |  |  |  | 7.15 | 10 | 0.711 |  |  |  |
| Married | 475 | 25.8% | 256 | 27.7% | 130 | 23.8% |  |  |  |  |  |  |
| Separated | 90 | 4.9% | 45 | 4.9% | 27 | 4.9% |  |  |  |  |  |  |
| Divorced | 170 | 9.2% | 85 | 9.2% | 46 | 8.4% |  |  |  |  |  |  |
| Widowed | 26 | 1.4% | 16 | 1.7% | 4 | 0.7% |  |  |  |  |  |  |

|  |  |  |  |  |  |  |
| --- | --- | --- | --- | --- | --- | --- |
| Never married | 1054 | 57.3% | 510 | 55.3% | 330 | 60.4% |
| Living with someone as married | 23 | 1.3% | 11 | 1.2% | 9 | 1.6% |

**Table 2.**
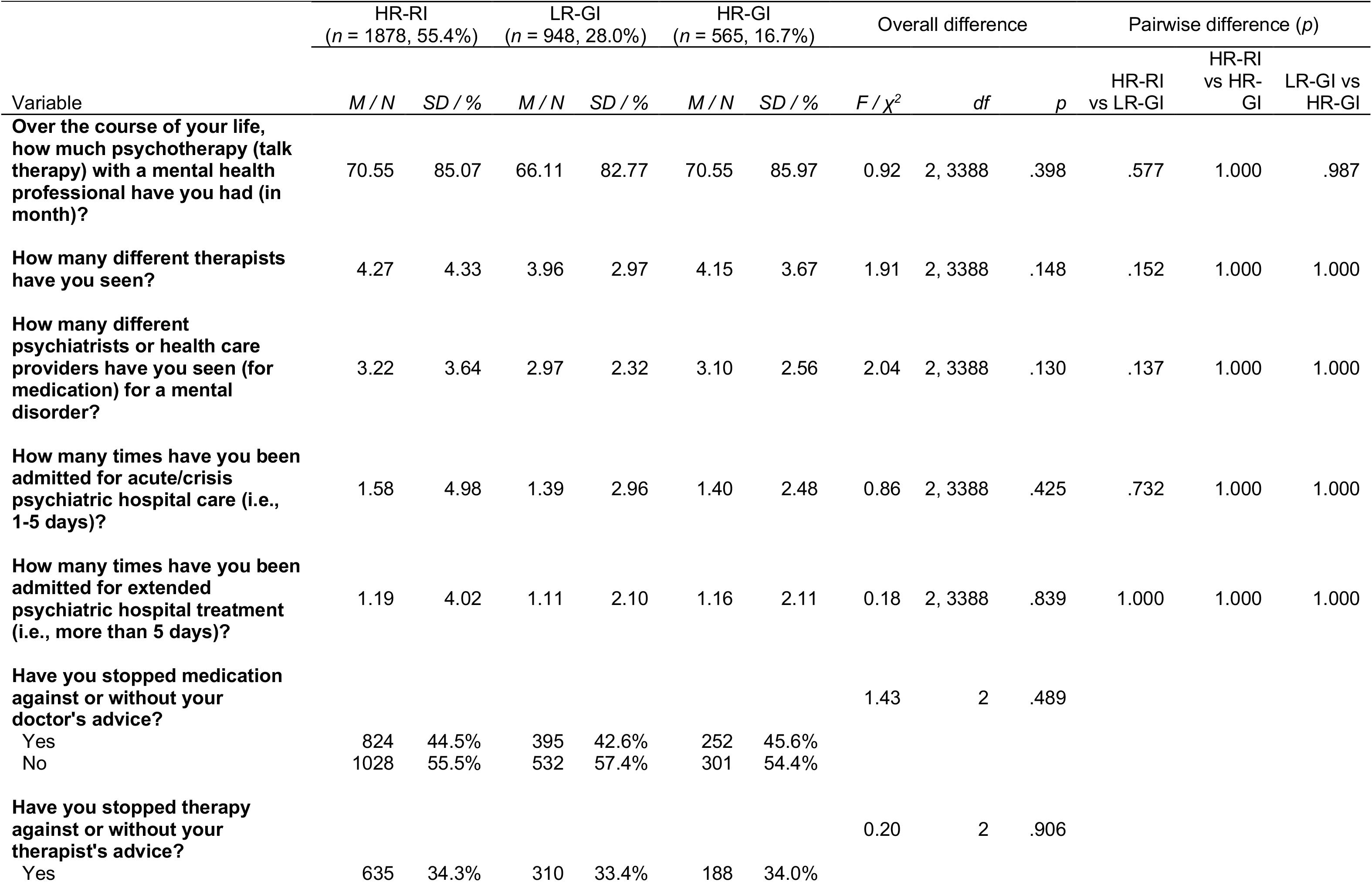
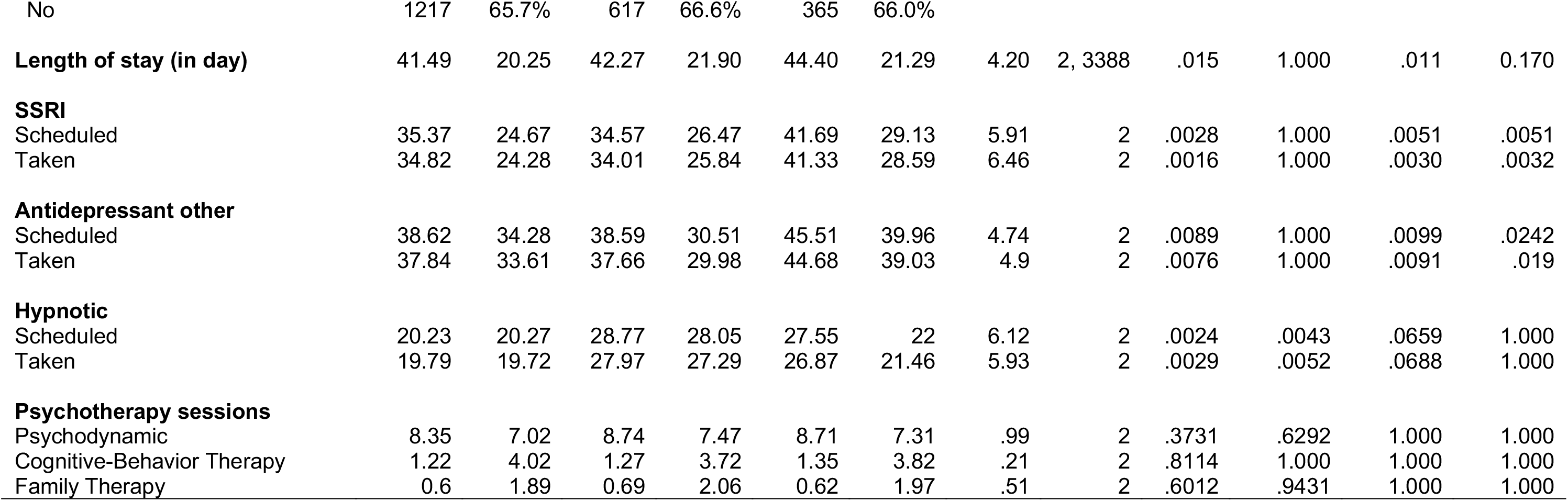
Comparisons of treatment variables and length of stay between the classes. HR-RI: High-risk, Rapid improvement, LR-GI: Low-risk, Gradual improvement, HR-GI: High-risk, Gradual improvement, M: mean, SD: standard deviation, SSRI: Selective Serotonin Reuptake Inhibitors.

| Variable | HR-RI<br>(n = 1878, 55.4%) |  | LR-GI<br>(n = 948, 28.0%) |  | HR-GI<br>(n = 565, 16.7%) |  | Overall difference |  |  | Pairwise difference (p) |  |  |
| --- | --- | --- | --- | --- | --- | --- | --- | --- | --- | --- | --- | --- |
| | M / N | SD / % | M / N | SD / % | M / N | SD / % | F / $\chi^2$ | df | p | HR-RI<br>vs LR-GI | HR-RI<br>vs HR-GI | LR-GI vs<br>HR-GI |
| **Over the course of your life, how much psychotherapy (talk therapy) with a mental health professional have you had (in month)?** | 70.55 | 85.07 | 66.11 | 82.77 | 70.55 | 85.97 | 0.92 | 2, 3388 | .398 | .577 | 1.000 | .987 |
| **How many different therapists have you seen?** | 4.27 | 4.33 | 3.96 | 2.97 | 4.15 | 3.67 | 1.91 | 2, 3388 | .148 | .152 | 1.000 | 1.000 |
| **How many different psychiatrists or health care providers have you seen (for medication) for a mental disorder?** | 3.22 | 3.64 | 2.97 | 2.32 | 3.10 | 2.56 | 2.04 | 2, 3388 | .130 | .137 | 1.000 | 1.000 |
| **How many times have you been admitted for acute/crisis psychiatric hospital care (i.e., 1-5 days)?** | 1.58 | 4.98 | 1.39 | 2.96 | 1.40 | 2.48 | 0.86 | 2, 3388 | .425 | .732 | 1.000 | 1.000 |
| **How many times have you been admitted for extended psychiatric hospital treatment (i.e., more than 5 days)?** | 1.19 | 4.02 | 1.11 | 2.10 | 1.16 | 2.11 | 0.18 | 2, 3388 | .839 | 1.000 | 1.000 | 1.000 |
| **Have you stopped medication against or without your doctor's advice?** |  |  |  |  |  |  | 1.43 | 2 | .489 |  |  |  |
| Yes | 824 | 44.5% | 395 | 42.6% | 252 | 45.6% |  |  |  |  |  |  |
| No | 1028 | 55.5% | 532 | 57.4% | 301 | 54.4% |  |  |  |  |  |  |
| **Have you stopped therapy against or without your therapist's advice?** |  |  |  |  |  |  | 0.20 | 2 | .906 |  |  |  |
| Yes | 635 | 34.3% | 310 | 33.4% | 188 | 34.0% |  |  |  |  |  |  |
| No | 1217 | 65.7% | 617 | 66.6% | 365 | 66.0% |  |  |  |  |  |  |
| **Length of stay (in day)** | 41.49 | 20.25 | 42.27 | 21.90 | 44.40 | 21.29 | 4.20 | 2, 3388 | .015 | 1.000 | .011 | 0.170 |
| **SSRI** |  |  |  |  |  |  |  |  |  |  |  |  |
| Scheduled | 35.37 | 24.67 | 34.57 | 26.47 | 41.69 | 29.13 | 5.91 | 2 | .0028 | 1.000 | .0051 | .0051 |
| Taken | 34.82 | 24.28 | 34.01 | 25.84 | 41.33 | 28.59 | 6.46 | 2 | .0016 | 1.000 | .0030 | .0032 |
| **Antidepressant other** |  |  |  |  |  |  |  |  |  |  |  |  |
| Scheduled | 38.62 | 34.28 | 38.59 | 30.51 | 45.51 | 39.96 | 4.74 | 2 | .0089 | 1.000 | .0099 | .0242 |
| Taken | 37.84 | 33.61 | 37.66 | 29.98 | 44.68 | 39.03 | 4.9 | 2 | .0076 | 1.000 | .0091 | .019 |
| **Hypnotic** |  |  |  |  |  |  |  |  |  |  |  |  |
| Scheduled | 20.23 | 20.27 | 28.77 | 28.05 | 27.55 | 22 | 6.12 | 2 | .0024 | .0043 | .0659 | 1.000 |
| Taken | 19.79 | 19.72 | 27.97 | 27.29 | 26.87 | 21.46 | 5.93 | 2 | .0029 | .0052 | .0688 | 1.000 |
| **Psychotherapy sessions** |  |  |  |  |  |  |  |  |  |  |  |  |
| Psychodynamic | 8.35 | 7.02 | 8.74 | 7.47 | 8.71 | 7.31 | .99 | 2 | .3731 | .6292 | 1.000 | 1.000 |
| Cognitive-Behavior Therapy | 1.22 | 4.02 | 1.27 | 3.72 | 1.35 | 3.82 | .21 | 2 | .8114 | 1.000 | 1.000 | 1.000 |
| Family Therapy | 0.6 | 1.89 | 0.69 | 2.06 | 0.62 | 1.97 | .51 | 2 | .6012 | .9431 | 1.000 | 1.000 |

## Materials and methods

### Participants

Study participants (*N* = 3,406) were adults admitted to a private inpatient psychiatric hospital in Houston, Texas between 2012 and 2017. The patients were equally split by sex (female: *n* = 1,637, 48.1%). On average, the patients were 35.24 years old (*SD* = 15.04; range = 17-89); and the majority of patients identified as White (*n* = 3,029, 88.9%) followed by multiracial or “other” (*n* = 176, 5.2%) and Asian (*n* = 71, 2.1%). LOS was defined as the number of days between admission date and discharge date for each participant. The average LOS of the participants was 42.16 days (*SD* = 20.9; range = 0-238 days). About half of the patients (*n* = 1,653, 48.5%) stayed at least six weeks in the hospital.

### Data procedures and measures

Data were collected as part of the clinic’s outcomes study aimed to monitor longitudinal treatment response during inpatient psychiatric treatment (Allen et al., 2009). We administrated clinical measures at admission, bi-weekly, and discharge. The study protocols were approved by the Institutional Review Board at Baylor College of Medicine.

The severity of depressive symptoms, anxiety symptoms, psychological flexibility, emotion regulation and dysregulation, and disability were measured by using the Patient Health Questionnaire (PHQ-9) (Kroenke et al., 2001), General Anxiety Disorder Scale (GAD-7) (Spitzer et al., 2006), Acceptance and Action Questionnaire II (AAQ-II) (Bond et al., 2011; Hayes et al., 2004), Difficulties in Emotion Regulation Scale (DERS) (Gratz and Roemer, 2004), and WHO Disability Assessment Schedule 2.0 (WHODAS 2.0) (Üstün et al., 2010), respectively. The Columbia-Suicide Severity Rating Scale (C-SSRS) was also used to examine suicidal ideation and behavior in the past 30 days and at present (Posner et al., 2011).

The Structural Clinical Interview for DSM-5 Disorders (SCID-5, SCID-5-PD) is the most widely used semi-structured clinical interview administered by a trained interviewer (First et al., 2015; First, 1997). The SCID-5-PD is used to assess for the presence of DSM-5 personality disorders. The SCID-5 is a diagnostic measure assessing mood disorders, psychotic disorders, substance use disorders, anxiety disorders, obsessive-compulsive disorders, eating disorders, somatic symptom disorders, insomnia, and trauma-related disorders. The SCID-5 and SCID-5-PD were administered at admission by trained clinical interviewers under supervision of a licensed psychologist.

As we have done previously (Hartwig et al., 2019; Oh et al., 2020), medication and psychotherapy information were extracted from patient medical records and treatment notes, respectively. Medication information (e.g., medication type, medication taken during the treatment) was measured one week prior to patient discharge date, as patients’ medication regimen had been stabilized by this time point. Medications were classified into the following categories: tricyclic antidepressants (TCA), selective serotonin reuptake inhibitors (SSRI), antidepressant other (e.g., Wellbutrin, trazodone), serotonin and norepinephrine reuptake inhibitors (SNRI), benzodiazepine, dopaminergic stimulants agents, first- and second-generation antipsychotics, hypnotics, non-opioid analgesics, and miscellaneous analgesics (e.g., gabapentin). Psychotherapy information (e.g., type of psychotherapy and frequency) was recorded in the patient’s treatment notes after receiving psychotherapies. Psychotherapies were classified into the following categories: psychodynamic, cognitive-behavioral therapy (CBT), and family therapy.

### Data analysis

GMM was used to classify patients into distinct classes, each manifesting a unique pattern of change in mental illness measured by six indicators: AAQ, DERS, GAD-7, PHQ-9, C-SSRS, and WHO-DAS. GMM postulates that similarities and differences in longitudinal observations may be explained by the existence of a categorical latent variable that represents a few mutually exclusive classes within the population (Nylund et al., 2007). In this study, a series of third-order GMM models were fitted to the patents’ scores on the six mental illness indicators observed at four different time points (admission, 2-week, 4-week, and 6-week; Fig. 1). Those six mental illness indicators were loaded on a first-order latent variable, named Mental Illness, at each time point; and the loadings of each indicator were constrained to be equal across time to achieve temporal measurement invariance. Cross-time residual covariances were also specified for each indicator in the models (they are omitted in Fig. 1 for the sake of simplicity). The Mental Illness latent variables were loaded on two or three second-order growth factors: Intercept and Linear Slope (2 latent variables); or Intercept, Linear Slope and Quadratic Slope (3 latent variables). Those growth factors were then loaded on a third-order class factor (1 categorical latent variable). A total of eight models were fitted – 1 to 4 unique patterns of linear growth (4 models) and another 1 to 4 unique patterns of quadratic growth (4 models) (Table 3). Model parameters, including factor loadings, means and variances/covariances of the growth factors, and class probabilities, were estimated by using robust maximum likelihood (MLR) via accelerated expectation-maximization algorithm, which often yields estimates and sandwich standard errors that are robust to non-normality and non-independence of data.

**Table 3.** Optimal number of growth patterns (classes). LL: log-likelihood, BIC: Bayesian information criterion.

| Model |  | LL | # parameters | BIC | Entropy |
| --- | --- | --- | --- | --- | --- |
| **Linear growth** | 1-class | −199336 | 78 | 399306 | - |
|  | 2-class | −195033 | 81 | 390724 | 1.000 |
|  | 3-class | −194920 | 84 | 390522 | 0.781 |
|  | 4-class | −194873 | 87 | 389921 | 0.720 |
| **Quadratic growth** | 1-class | −199130 | 82 | 398927 | - |
|  | 2-class | −194674 | 86 | 390047 | 0.999 |
|  | 3-class | −194509 | 90 | 389750 | 0.796 |
|  | 4-class | −194412 | 94 | 389588 | 0.685 |

**Fig 1.**
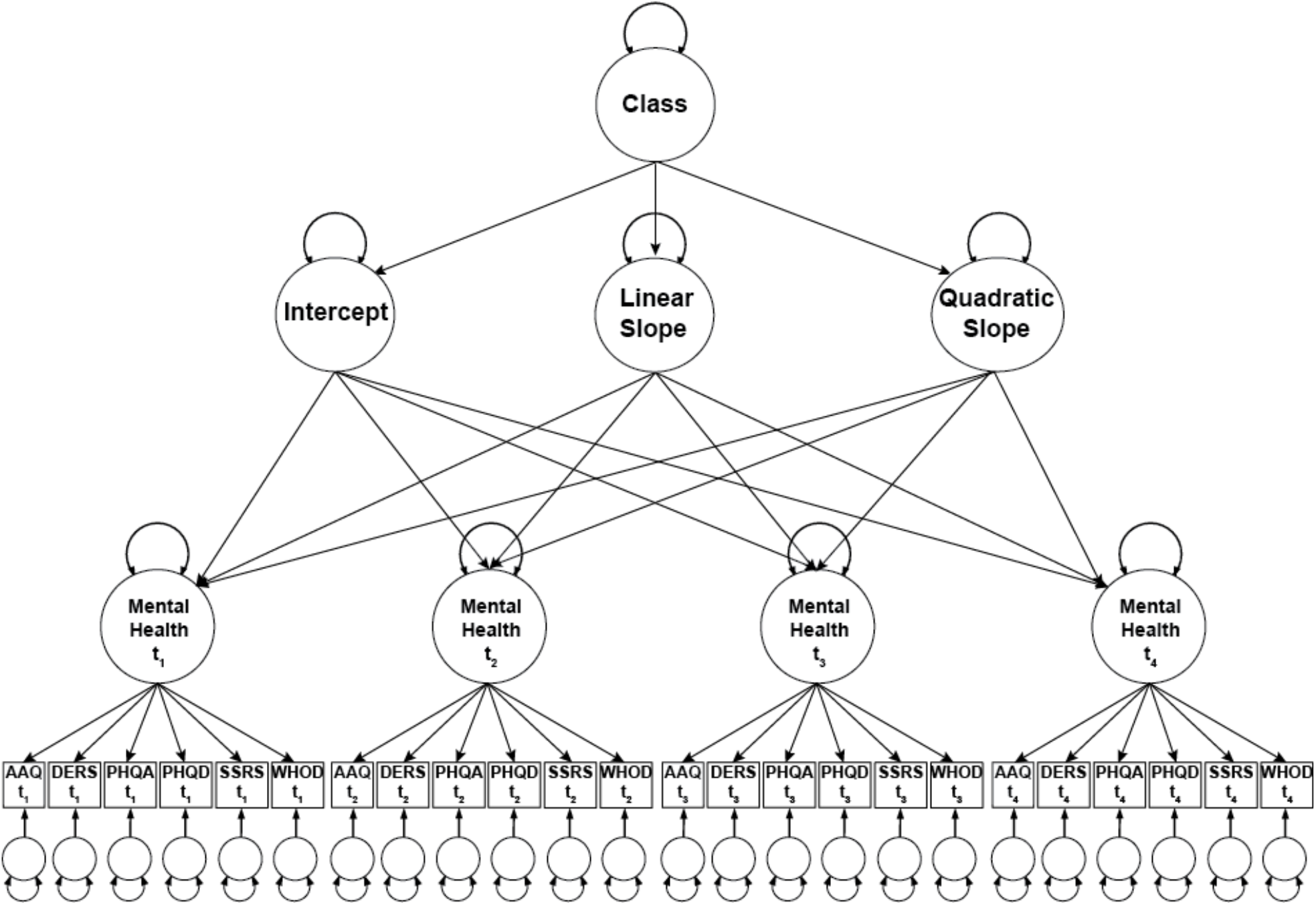
An example of fitted growth mixture models (GMM). t1: admission, t2: week 2, t3: week 4, t4: week6, AAQ: psychological flexibility, DERS: emotion regulation problem, PHQA: anxiety, PHQD: depression, SSRS: suicide ideation severity, WHOD: disability.

Next, the shape of growth (linear vs. quadratic) and the optimal number of growth patterns (classes) were determined by comparing the models in terms of (a) entropy that quantifies the amount of classification error (Celeux and Soromenho, 1996; Ramaswamy et al., 1993) – a larger value and close to 1 indicates less classification error made by the model; (b) average classification posterior probabilities (ACPP) (Nagin, 1999) – high values at the diagonal of a classification table and low values at the off-diagonal of the table indicate good classification quality; and (c) Bayesian Information Criterion (BIC) (Sclove, 1987) – lower values indicate a better fitting model. Note that Parametric bootstrap likelihood-ratio test (BLRT) (McLachlan et al., 2000) was also performed, but the test results were discarded because they had local maxima issues in the bootstrapping process. Entropy and ACPP could not be computed for the case of single class and thus comparisons were not made against the 1-class models. Once the GMM analysis successfully identified distinct classes of patients who share similar longitudinal trajectories of mental illness, a post-hoc analysis was conducted to understand the characteristics of these classes – or equivalently, to find potential antecedents and consequences of differential treatment efficacy. In this second stage, the identified classes were compared in terms of theoretically relevant and meaningful variables such as age, sex, ethnicity, marital status, educational level, occupation, previous experiences of psychotherapy, hospital care, medication use history, and current length of stay, etc. Chi-square tests of independence were used for the comparisons of categorical variables; independent-samples t-tests or analyses of variance (ANOVA), depending on the number of identified classes, were performed for the comparisons of continuous variables. Statistical significance was determined at a 0.05 alpha level, or at a level reduced to control for Type I error in multiple pairwise comparisons (i.e., Bonferroni adjustment). All analyses were conducted using Mplus 8.0 (Muthén and Muthén, 2018) and SAS 9.4 (SAS Institute, 2013).

## Results

### Find the distinct patterns of recovery in mental illness

The patients were grouped into three mutually exclusive classes as optimally representing three different patterns of (quadratic) change in mental illness over the inpatient treatment period. Specifically, the entropy values of greater than .80 showed a clear delineation of classes in the ‘quadratic’ GMM models with two or three classes (Celeux and Soromenho, 1996). Further, the ACPP values from the ‘quadratic’ model with three classes (.84-.92 at the diagonal of the classification table and 0-.08 at the off-diagonal) denoted the best classification quality among the models being tested. The BIC values decreased as the number of classes increased in the case of either linear or quadratic patterns of growth. However, caution should be exercised when interpreting this result because BIC tends to overestimate the number of classes (Enders and Tofighi, 2008; Henson et al., 2007; Nylund et al., 2007). Thus, considering the results of entropy and ACPP, we chose the ‘quadratic’ GMM model with three classes as the final solution.

The final model solution was substantively interpretable – the identified classes of patients indeed represented three distinctive patterns of change in mental illness (see Fig. 2). The first class, the largest class including 55.4% of the sample, showed more severe mental illness at the beginning of the treatment (i.e., high latent scores) but their mental illness improved substantially across inpatient psychiatric treatment, particularly in the first two weeks of treatment. Thus, this class was referred to as High-Risk, Rapid Improvement (HR-RI). The second largest class (28%) included patients with less severe mental illness at the start of the treatment (i.e., low latent scores) who made gradual improvement during the treatment. The extent of recovery was lessened at the later stage of the treatment (i.e., the slope of the line is closer to 0). This second class was referred to as Low-Risk, Gradual Improvement (LR-GI). The last class of patients (16.7%) was characterized by more severe mental illness at the beginning of the treatment, similar to the HR-RI class, but these patients made only gradual improvements over the treatment period – High-Risk, Gradual Improvement (HR-GI). The means of the individual indicators of mental illness and their changes at four different time points are shown in Fig. 3. Additionally, follow-up assessments of anxiety and depressive symptoms (at 2 weeks, 3 months, 6 months, and 1-year post-discharge) are displayed in Fig. 3A. As expected, all observed scores were well aligned with the ‘model-implied’ longitudinal trajectories (i.e., estimated latent scores) of mental illness during inpatient psychiatric treatment.

**Fig 2.**
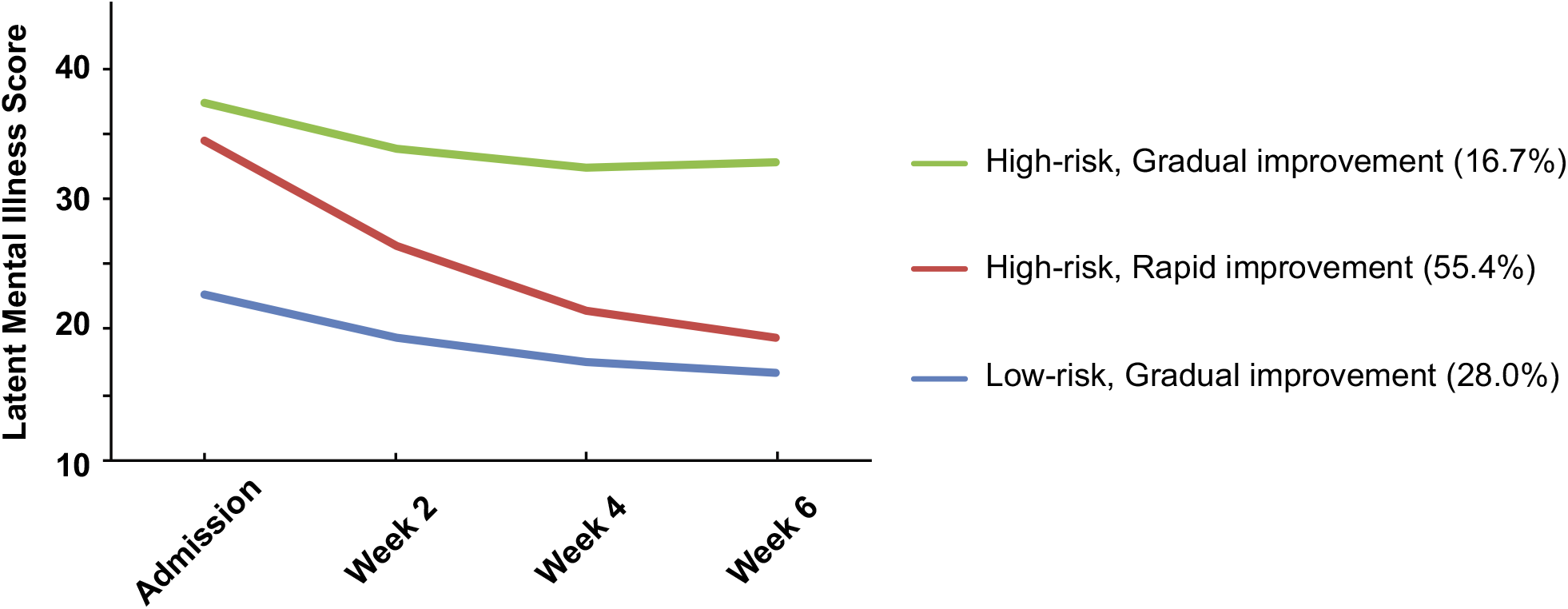
Latent mental illness score in identified classes. Higher latent mental illness scores indicate more disabled mental illness.

**Fig 3.**
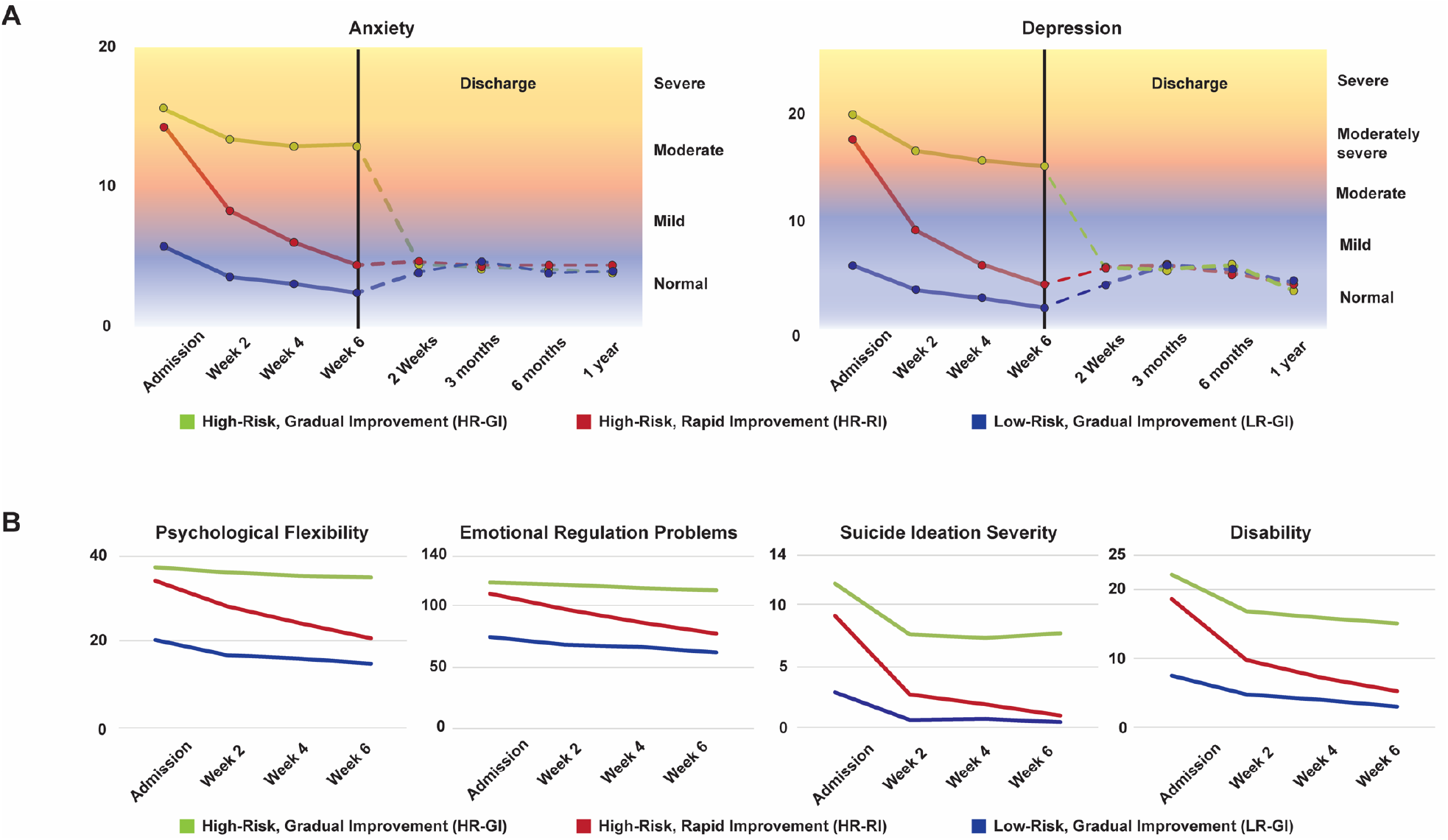
Observed scores of six indicators of mental illness. A. Each number at Anxiety and Depression indicates the mean of PHQANXIETY and PHQDEPRESSION, respectively. Follow-up assessments after discharge were characterized in the normal range. B. Each number at Psychological Flexibility, Emotional Regulation Problems, Suicide Ideation Severity and Disability indicates the mean of AAQ, DERS, SSRS and WHOD scores, respectively.

### Identify patient characteristics associated with differential treatment efficacy

The post-hoc analyses revealed similarities and differences among the three identified classes. Table 1 and 2 provide the descriptive statistics, the results of bivariate tests, ANOVA, and follow-up pairwise comparisons. On average, the HR-GI class (33.24 ± 13.71) was significantly younger than both the HR-RI (35.26 ± 14.99; adjusted *p* < 0.05) and the LR-GI (36.38± 15.79; adjusted p < 0.001) classes, while the age of the latter two classes was not significantly different (adjusted *p* = 0.18). The LR-GI class had more male patients (63.4%) than females (36.6%), while the HR-GI class had more female patients (59.8%) than males (40.2%) (*p* < 0.001); The HR-RI class had an equivalent number of males (49.6%) and females (50.4%). The three classes demonstrated similar ethnic breakdowns (*p* = 0.33) – the majority of the class members were white (87.9-90.1%) followed by multiracial or “other” category (5-5.2%) and Asian (1.5-2.3%), as observed in the overall sample. The patients’ marital status did not differ among the three classes (*p* = 0.71). Also, the patients’ education level (*p* = 0.23) and occupation (*p* = 0.97; see Supplementary) were not significantly related to their patterns of recovery.

Regarding clinical history and LOS in treatment, the patients in three different classes reported similar previous experiences of psychotherapy, hospital care, and stopping medication (all *p* > 0.05; see Table 2). However, significantly fewer patients in the HR-RI class took hypnotic medications (19.79 ± 19.72%) compared to the LR-GI class (27.97 ± 27.29%) (adjusted *p* < .01). Also, significantly fewer patients in the HR-RI class took antidepressant other (e.g., Wellbutrin, trazodone) (37.84 ± 33.61%) and SSRI (34.82 ± 24.28%) medications compared to the HR-GI class (44.68 ± 39.03% and 41.33 ±28.59%, respectively) (both adjusted *p* < .01). The HR-RI class (41.49 ± 20.25) stayed in their current hospitalization for significantly shorter periods of time than the HR-GI class (44.40 ± 21.29; adjusted *p* < 0.05), with the HR-RI group staying approximately 3 days shorter than the HR-GI class. The average LOS for the LR-GI class (42.27 ± 21.90) was in between the average LOS for the two High-Risk classes and did not significantly deviate (adjusted *p* = 1.00 and 0.17). SCID-5 and SCID-5-PD DSM-5 clinical diagnosis differences were also examined within each class (see Supplementary). The most common diagnoses in both the HR-RI and LR-GI classes were (1) major depressive disorder, recurrent (HR-RI = 42%; LR-GI = 39%), followed by (2) anxiety not otherwise specified (HR-RI = 27%; LR-GI = 31%), (3) substance dependence disorder (HR-RI = 27%; LR-GI = 29%), (4) generalized anxiety disorder (HR-RI = 19%; LR-GI = 20%), and (5) alcohol abuse disorder (HR-RI = 17%; LR-GI = 16%). The HR-GI class also had similar common diagnoses to the HR-RI and LR-GI classes, but with substance dependence disorder more prevalent than anxiety not otherwise specified: (1) major depressive disorder, recurrent (HR-GI = 45%), followed by (2) substance dependence disorder (HR-GI = 28%), (3) anxiety not otherwise specified (HR-GI = 26%), (4) generalized anxiety disorder (HR-GI = 21%), and (5) alcohol abuse disorder (HR-GI = 16%). Finally, the type and frequency of psychotherapy sessions received during inpatient psychiatric treatment did not differ between the three classes (*p* = .37; see Table 2).

## Discussion

Aims of the current study were to (a) find the distinct classes of patients who demonstrate similar trajectories of mental illness outcomes during inpatient psychiatric treatment, and (b) identify patient characteristics associated with the identified patterns of recovery over time. Through the use of growth mixture modeling, we were able to describe three distinct trajectories (HR-RI, LR-GI, and HR-GI) of mental illness improvement across inpatient psychiatric hospitalization as measured by six indicators: psychological flexibility, emotion regulation problems, anxiety, depression, suicide ideation severity, and disability. Patients’ age and sex were significantly related to treatment response, with significantly younger females in the HR-GI group and more males in the LR-GI group.

The findings of the present study support our hypothesis that different mental illness trajectories would emerge by identifying three classes of patients that are distinct in terms of mental illness at the beginning of treatment and change over their hospitalization. These findings suggest that the patient classes could be differentiated at the beginning of the treatment and by the improvement of six indicators during inpatient psychiatric treatment. The most common trajectory class was characterized by more severe mental illness at the beginning of the treatment and rapid treatment improvement during inpatient psychiatric treatment, referred as HR-RI (55.4%). The second class, LR-GI (28%), was characterized by less severe mental illness than the HR-RI group at the start of the treatment and gradual improvement during the inpatient treatment. The last class of patients, HR-GI (16.6%), demonstrated the highest severity of mental illness at admission and gradual improvement over the treatment period. Thus, the majority of patients could be categorized as following a path to recovery from a fairly severe experience of mental health and only a smallish group (16%) of severely impacted patients could be categorized as poor responders.

Having said that, all trajectory classes demonstrated improvement (significant negative slope of trajectories) in mental illness across treatment and their improvement was most rapid after psychiatric hospitalization, consistent with earlier studies that reported depressive symptoms and suicide ideation reduction following the first week of admission or even brief intervention (Clapp et al., 2013; Czyz and King, 2015; Hirsch et al., 1979; Hopko et al., 2001; Lieberman et al., 1998; Pettit et al., 2005; Prinstein et al., 2008; Rocca et al., 2010). Factors contributing to rapid improvement may include medication effects (e.g., antidepressant, antipsychotic agents) (Agid et al., 2003; Posternak and Zimmerman, 2005), removal of alcohol and substances, and psychotherapy gains (Stiles et al., 2003). All classes sustained their improvement related to anxiety and depression post-discharge from the hospital (i.e., anxiety and depression scores post-discharge were in the normal range (Kroenke and Spitzer, 2002; Spitzer et al., 2006)), although generalization of these findings is limited as there was participation attrition at follow-up.

Our results are consistent with other studies demonstrating the follow-up assessments of depressive symptoms after hospitalization for traumatic brain injury (Bombardier et al., 2016) and maternal depressive symptoms from pregnancy through 2 years postpartum (Mora et al., 2008). However, these prior studies also reported a trajectory showing gradual reemergence of depressive symptoms which may associate with a history of alcohol dependence and other mental illness disorders (Bombardier et al., 2016). In the current study, we found that patients’ age and sex best predict the particular patterns of mental illness recovery trajectories. The HR-GI class (33.24 ± 13.71) had patients who were significantly younger relative to the HR-RI (35.26 ± 14.99) and LR-GI (36.38 ± 15.79) classes. The LR-GI class had fewer female patients than males, whereas HR-GI class had more female patients than males. These findings are consistent with previous studies that females have shown higher rates of depressive symptoms (Kandel and Davies, 1982), generalized anxiety disorder (Altemus et al., 2014; Kessler et al., 1994), and suicidal ideation (Borges et al., 2006; Crosby et al., 1999). Additionally, previous studies have consistently demonstrated higher rates of depressive symptoms (Kessler et al., 2010) and suicidal ideation (Crosby et al., 1999) among young adults. For the LR-GI class that had more men than women, as they were admitted to an inpatient psychiatric hospital, the likelihood that they did not present with mental illness symptoms is unlikely. Instead, as men have a higher rate of minimizing self-reported symptoms due to societal ideals of masculinity (Martin et al., 2013), it is possible that symptoms are minimized by this patient group.

We examined the LOS in each trajectory class and found that the HR-RI class stayed in the hospital less time (by a few days) than the HR-GI class (adjusted *p* < 0.05). Note that the average LOS in this study was approximately 42.16 days which is a contrast to the previous studies (five to six days being the average LOS) (Glick et al., 2011; Sturm and Bao, 2000) that indicate negative treatment outcomes, including higher rates of death by suicide post-discharge (Appleby et al., 1999; Goldacre et al., 1993). Therefore, our results highlight not only better understanding of the distinct classes but emphasize the importance of a longer LOS in generating positive treatment outcomes for inpatient psychiatry. Previous studies have shown that patients with cognitive impairment had a significantly increased LOS, but such differences in LOS were not found among patients with depression and anxiety (Furlanetto and da Silva, 2003). However, other studies have reported that depression, anxiety, schizophrenia, mood disorders, and alcohol and drug related disorders are important predictors of LOS (Borchardt and Garfinkel, 1991; Draper and Luscombe, 1998; Huntley et al., 1998; Jiménez et al., 2004; Sloan et al., 1999) and that psychiatric comorbidity could have contributed to prolonged LOS (Bressi et al., 2006; Saravay, 1994). In addition to clinical diagnoses, a previous study reported that longer LOS was associated with sex (i.e., females) (Averill et al., 2001). It should be noted that the most common diagnoses in all three classes were recurrent major depressive disorder, anxiety not otherwise specified, and substance dependence disorder. The HR-GI class, which showed longer LOS than the HR-RI class, had more female patients than males, as compared to other two classes. Clearly, the hypothesis (i.e., improved mental illness outcomes would be associated with longer LOS) was not supported by the data, but this group of patients deserves more careful scrutiny and their treatment path (more SSRI and more antidepressant other) indicates that during their slightly longer treatment a wider range of therapies were tried. Therefore, it is likely that significant factors related to LOS for inpatient psychiatry may relate to demographic factors, particularly sex. Perhaps, a bi-factor analytic approach would show them to have higher p Factor scores than the other two groups indicating a higher level of general psychopathology (Caspi and Moffitt, 2018; Smith et al., 2020).

A number of limitations of the present study should be addressed. First, the study sample was from an inpatient psychiatric hospital where most patients were white and had a higher level of education than the general population. Therefore, a future study needs to be replicated with a more diverse sample in order to ensure our findings are not limited to the present sample. Although follow-up assessments of depression and anxiety showed that patients in this study maintained their improved status throughout their first year after discharge, the follow-up results should be interpreted with caution because there is potential for bias in the missing assessments (e.g., individuals who were in other treatment facilities post-discharge) - missing data after discharge: 82.8% at 2 weeks, 85.2% at 3 months, 88.3% at 6 months, and 91.1% at 1 year. While obtaining a more comprehensive follow up sample will always be challenging in the context of routine outcomes monitoring, data obtained from such a small proportion of the sample is very likely to be irredeemably contaminated by non-random forces associated with loss to follow-up (such as an understandable wish to obliterate an experience that may have felt stigmatizing). Data with fewer missing and frequent follow-up assessments of the indicators used in this study may help to improve our understanding of distinct trajectory classes after discharge and how clinical outcomes are/are not maintained following post-discharge.

In this study, we identified three distinct mental illness trajectory classes using a large sample of 3,406 patients admitted to a private inpatient psychiatric hospital. The majority of the patients with high mental illness at the beginning experienced substantial improvement within 6 weeks of inpatient psychiatric treatment and maintained their reduction in depression and anxiety throughout their first year after discharge. This study highlights the importance of understanding the relevance of the distinction of classes with meaningful patterns of mental illness treatment change over time. The knowledge of differences between the classes may provide valuable information for the clinicians as well as future researchers making predictions regarding the course of mental illness improvement during inpatient psychiatric treatment and after hospitalization, and ultimately to ensure that patients receive the adequate length of stay for inpatient psychiatric treatment in order to maximize their mental illness outcomes.

## Data Availability

The data that support the findings of this study are available on request from the corresponding author. The data are not publicly available due to [restrictions e.g. their containing information that could compromise the privacy of research participants].

## Acknowledgement

We thank past and present members of the research team at The Menninger Clinic for their contributions to research and data collection. We also thank The Menninger Clinic Information Technology department who provide the technical support to conduct research effectively.

## Contributors

All authors have approved the final article. Conception: HO, JL, KR, JO, BS, and MP. Data analysis: HO, JL, SK, KR, and MP. Interpretation: HO, JL, KR, PF, JO, and MP. Drafting the manuscript: HO, JL, KR, and MP.

